# Multiple hypervirulent methicillin-sensitive *Staphylococcus aureus* lineages contribute towards poor patient outcomes in orthopedic device-related infections

**DOI:** 10.1101/2022.10.21.22280349

**Authors:** Virginia Post, Ben Pascoe, Evangelos Mourkas, Jessica K. Calland, Matthew D. Hitchings, Christoph Erichsen, Julian Fischer, Mario Morgenstern, R. Geoff Richards, Samuel K. Sheppard, T. Fintan Moriarty

**Affiliations:** AO Research Institute Davos, Davos, Switzerland; Centre for Genomic Pathogen Surveillance, Big Data Institute, University of Oxford, Oxford, United Kingdom; Ineos Oxford Institute of Antimicrobial Research, Department of Biology, University of Oxford, Oxford, United Kingdom; Faculty of Veterinary Medicine, Chiang Mai University, Chiang Mai, Thailand; Oslo Centre for Biostatistics and Epidemiology, Oslo University Hospital, Oslo, Norway; Swansea University Medical School, Swansea University, Swansea, United Kingdom; Department of Trauma Surgery, Trauma Centre Murnau, Murnau, Germany; Department of Orthopedic and Trauma Surgery, University Hospital Basel, Switzerland

**Keywords:** *Staphylococcus aureus*, MRSA, MSSA, virulence factors, antibiotic resistance, orthopedic device-related infections

## Abstract

Staphylococci are the most common cause of orthopedic device-related infections (ODRIs), with *Staphylococcus aureus* responsible for a third or more of cases. This prospective clinical and laboratory study investigated the association of genomic and phenotypic variation with treatment outcomes in ODRI isolates. Eighty-six invasive *S. aureus* isolates were collected from patients with ODRI, and clinical outcome was assessed after a follow-up examination of 24 months. Each patient was then considered to have been “cured” or “not cured” based on predefined clinical criteria. Whole genome sequencing and molecular characterization identified isolates belonging to globally circulating community- and hospital-acquired pandemic lineages. Most isolates were phenotypically susceptible to methicillin and lacked the SCC*mec* cassette (MSSA), but contained several (hyper) virulence genes, including toxins and biofilm genes. While recognizing the role of the host immune response, we identify characteristics of isolate genomes that, with larger datasets, could help contribute to infection severity or clinical outcome predictions. While this and several other studies reinforce the role antibiotic resistance (e.g., MRSA infection) has on treatment failure, it is important not to overlook MSSA that can cause equally destructive infections and lead to poor patient outcomes.

**Importance:** *Staphylococcus aureus* is a prominent cause of orthopedic device-associated infections, yet little is known about how the infecting pathogen, and specifically the repertoire of genome-encoded virulence factors can impact treatment outcome. Past studies have focused on distinguishing commensal from invasive *S. aureus* isolates but in this study, we aim to investigate traits in infecting isolates that influence patient outcomes. Invasive *S. aureus* isolates were collected from orthopedic-device related infection patients and categorized according to the success of subsequent treatment (“cured” /”not cured”), as determined following hospital discharge two years after initial presentation. Several MSSA hypervirulent clones were associated with a “not cured” clinical outcome. Improved understanding of the bacterial traits associated with treatment failure in ODRI will inform the risk assessment, prognosis, and therapy of these infections.

## Introduction

The most challenging complication in orthopedic surgery is orthopedic-device related infection (ODRI), with incidence ranging from 0.7 % to 4.2 % for elective orthopedic surgeries (1–4). Incidence increases to over 30 % following operative fixation of complex open fractures (5, 6). While patient health is a key risk factor (including a high BMI and chronic immunosuppression) for poor treatment outcomes (7, 8), there is evidence that pathogen genetic diversity can be an indicator of patient outcome (9–12). *Staphylococcus aureus* is the most common infecting agent (2, 3, 13–15) and treatment outcome is often complicated by infection with antimicrobial resistant lineages e.g., methicillin-resistant *S. aureus* (MRSA) (8, 16). Combined with high virulence potential, MRSA are difficult to treat and are a global healthcare concern. Despite this, predicting treatment outcomes based on bacterial phenotypes/genotypes remains difficult (8, 10, 11, 17).

Hypervirulent pandemic clonal lineages have helped spread *S. aureus* around the world and the geographic distribution of many lineages is dynamic, with different lineages dominating infections in specific global regions (18, 19). Waves of MRSA lineages have risen and been replaced since the emergence of MRSA in the 1940s (18, 20). These highly structured populations can be grouped into clonal complexes (CCs) that share five or more alleles at seven multi-locus sequence typing (MLST) loci (21–23). Community-associated MRSA (CA-MRSA) have begun to replace hospital-associated MRSA (HA-MRSA) as the dominant epidemic strains (24). The most prevalent lineages include 5 global community acquired (CA-MRSA) genotypes CC1, CC8, CC30, CC59 and CC80. The most common hospital-acquired lineages are CC5, CC22 (UK), CC239 and CC45 (21–23). Combining MLST with traditional molecular typing techniques such as identification of the staphylococcal cassette chromosome *mec* (SCC*mec*) (25) and *spa* repeat regions (26) can provide a nomenclature to describe relevant epidemic clones can be provided. Lineages can acquire advantageous traits, such as antibiotic resistance, which proliferate in the population through the decedents of successful strains. This likely occurs in many instances, however, the extent of this in the context of ODRI remains to be determined.

Despite the growing concern of MRSA lineages, MSSA isolates are often the most common in invasive surgery-related infections (27, 28). Convergent evolution in several CA-lineages potentially balances the fitness costs of expressing AMR genes with the acquisition of multiple virulence factors (29–32). Specific genes encoding putative virulence factors in invasive *S. aureus* disease involve evasion of immune defenses, including the ability to adhere to and invade host tissues - essential for ODRI (33, 34). A large body of work has identified many virulence factors, including Microbial Surface Components Recognizing Adhesive Matrix Molecules (MSCRAMMs), the polysaccharide intercellular adhesion (PIA), the Staphylococcal protein A, extracellular proteins such as coagulase, Staphylococcal enterotoxins (SE), exfoliatins (ET), toxic shock syndrome toxin (TSST), staphyloxanthin, hemolysins and Panton Valentine leukocidin (PVL) that have a crucial impact on the pathogenicity of *S. aureus* infections (35–38). Genome-wide association studies have been applied to numerous bacterial species (39, 40), and identified genes or genetic elements associated with disease that transcend clonal patterns of inheritance (not confined to specific lineages) in Staphylococci. Phenotype filtering techniques have attempted to assess patient risk and predict disease outcome from genetic data based on enrichment for disease-associated traits (12, 41–43). GWAS approaches have shown promising disease prediction results (31, 43, 44), as well as AMR profiles (45). As in other bacterial species (46–48), a better understanding of genome and transcriptome variation in hypervirulent infection types shows promise for our ability to predict disease severity in Staphylococci (49).

In this study we investigate the association between treatment outcomes determined 2 years post-operatively in patients with *S. aureus* ODRI and phenotypic and genotypic features of the infecting pathogen. Building on recent studies aiming to predict *S. aureus* virulence from genome sequence (43, 44), we aim to distinguish high risk lineages, isolates and genes. These features were correlated with mortality rate in a simplified virulence model using *Galleria mellonella*.

## Results

*S. aureus* isolates were collected from 86 patients undergoing operative revision of an ODRI (**Supplementary table S1**). Patient outcomes were assessed after a 2-year follow-up period and “cured” patients were free of infection, surgical and systemic antibiotic therapy had ceased with function of the affected joint or limb restored. If one or more of these parameters was negative, patients were considered to have had a “not cured” outcome (8, 12). Most patients enrolled in the study received successful treatment and were among the “cured” cohort (**Table 1**; n= 65/86; 75.6%). Treatment was unsuccessful in 21 patients (n=21/86; 24.4% “not-cured”) and multiple revision surgeries were necessary for nearly all patients (n=83/86; 96.5%).

**Table 1:** Cohort description

|  |  | **Complete study cohort** |  |
| --- | --- | --- | --- |
| **Total, n (%)** |  | **86** | **(100%)** |
| ***Clinical course and infection outcome, n (%)*** |  |  |  |
| Multiple revision surgeries |  | 83 | 96.5% |
| Cured |  | 65 | 75.6% |
| ***Health status, n (%)*** |  |  |  |
| Obesity |  | 25 | 29.1% |
| Smoking |  | 28 | 32.6% |
| Diabetes |  | 16 | 18.6% |
| Chronic immunosuppression |  | 11 | 12.8% |
| ***Infection characteristics, n (%)*** |  |  |  |
| Peri-prosthetic joint infection |  | 19 | 22.1% |
| Acute infection |  | 25 | 29.1% |
| Open fracture (initially) |  | 21 | 24.4% |
| ***Type of implant, n (%)*** |  |  |  |
| Peri-prosthetic joint infection (PJI) |  | 19 | 22.1% |
| Nail |  | 19 | 22.1% |
| Plate |  | 36 | 41.9% |
| Screw |  | 6 | 7.0% |
| Fixateur externe |  | 4 | 4.7% |
| K-wire |  | 1 | 1.2% |
| Ligament reconstruction device |  | 1 | 1.2% |

### Host-associated risk factors

Extensive patient data, types of implanted devices and clinical presentation were recorded for each infected patient (**Table 1**). Additionally, the effects of patient co-morbidities (such as diabetes or obesity), fracture types or the time of symptoms onset on treatment outcome were analyzed (**Table 2**). None of these prognostic factors alone significantly decreased cure rate.

**Table 2:**
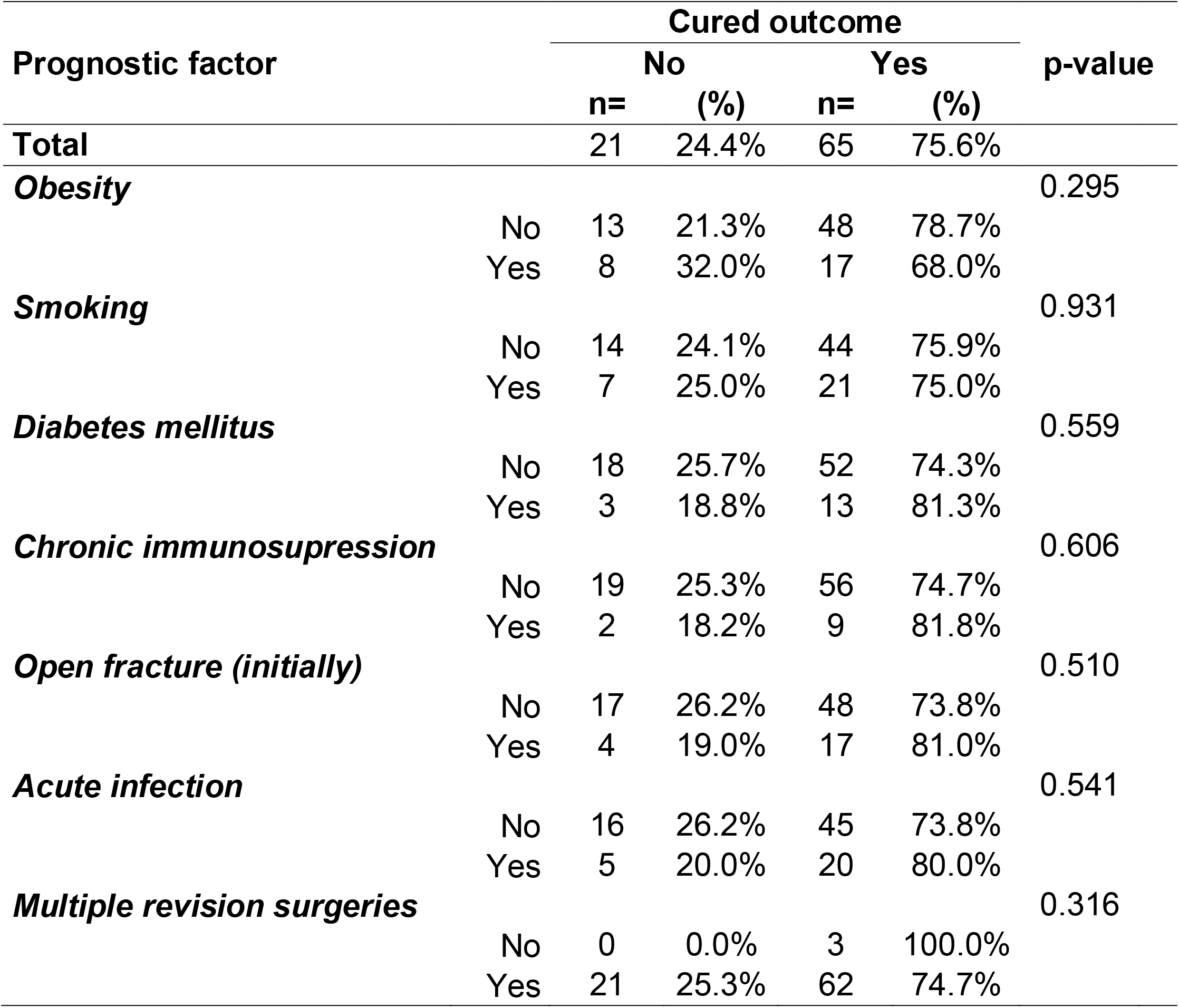
Patient risk factors

### Multiple lineages contribute to poor patient outcome

A maximum-likelihood phylogeny was constructed based on shared coding sequences, present in 95% or more isolates (**Figure 1A; Supplementary table S1**). The collected isolates were genetically diverse and represented 19 different sequence types (STs) (**Figure 1B**) based on the 7 loci scheme for *S. aureus* and could be grouped into 18 clonal complexes (CCs) based on 5 or more shared MLST loci (21). No clear clustering was observed between “cured” and “not-cured” isolates. Consistent with other European surveillance efforts, six of our seven most common lineages matched common pandemic *S. aureus* lineages (CC5, CC8, CC22, CC30, CC45 and CC59) previously identified by the ESCMID study group (collected from 26 European countries between 2006-7) (50). Our isolates predominantly lacked the *SCCmec* cassette, which confers methicillin resistance and were classified as methicillin-susceptible *S. aureus* (MSSA; n= 81/86; 94%) (**Figure 1C**).

**Figure 1:**
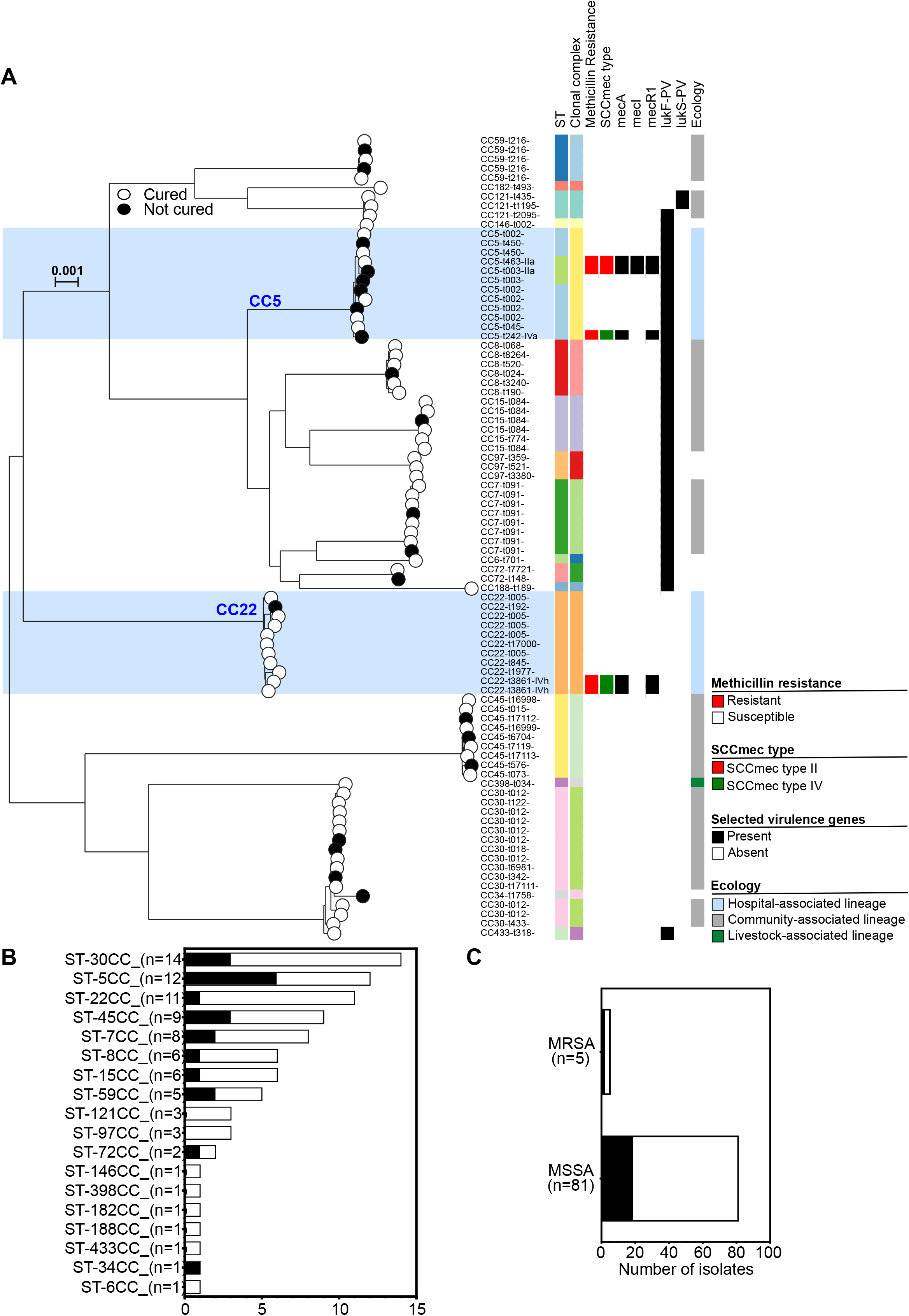
Population structure of *S. aureus* isolates collected in this study. **A:** A maximum-likelihood phylogeny was constructed with IQ-TREE, using a GTR model and ultrafast bootstrapping (1000 bootstraps; version 2.0.3)(108, 109) from an alignment of all isolates (n=86). Scale bar represents a genetic distance of 0.001. Leaves from isolates that were “cured” are white (n=65) and those that did not achieve a “cured” status are black (n=21). The tree is annotated with MLST, clonal complex, methicillin resistance status, *SCCmec* and *spa* types and the presence of PVL genes (indicated by colored blocks). Isolates from hospital associated lineages are highlighted in blue. Interactive visualization is available on Microreact (110): https://microreact.org/project/Post-Pascoe-Saureus/0f0c26fb. The number of isolates from each (**B**) MLST clonal complexes (CC) and (**C**) methicillin resistant (MRSA) and methicillin susceptible (MSSA) lineages. The proportion of isolates that lead to a “not cured” status is shown in black.

Most of our isolates (n=48/86; 55.8%) were from community-acquired lineages (MSSA or SCC*mec* types I, II, III), including CC30 which was the most common clonal complex identified in our collection (n=14/86; 16.3%) (**Figure 1A+B**). In more than three quarters (n=11/14; 78.6%,) of cases where this clonal complex was identified, the patient was deemed to have had a good outcome and a “cured” status (**Supplementary table S2**). A minority of isolates were identified from other CA-lineages, CC8 (n=6) and CC59 (n=5), CC45 (n=9), CC7 (n=8), CC15 (n=6). No isolates were sampled from two common pandemic CA-lineages, CC1 and CC80.

Many isolates we identified were from well described hospital-acquired lineages (n=23/86; 26.7%), including the globally distributed CC5 lineage (n=12) (**Figure 1A+B**). Infections from this CC were often unresolved (n=6/12; 50%) and posed the highest risk of a “not cured” patient outcome. CC22 is the most common sequence type identified from clinical infections, particularly in the UK (51, 52), but was the 3^rd^ most sampled CC in our collection (n=11/86; 12.8%). This clonal complex was implicated in a “not cured” patient outcome on only one occasion. All other CCs were represented by fewer than 5 isolates. One isolate was isolated from the livestock associated lineage, CC398 (n=1).

#### Accessory genome differences in ODRI isolates

ODRI that leads to a “not cured” patient outcome is a complex process, which is affected by genetic and environmental differences in the host (**Table 2**) as well as variation in the infecting bacterial population. To investigate differences in gene presence between isolates showing phenotypic variation, we further investigated genes in the accessory genome. We characterized the pangenome using PIRATE (53) with 86 isolates plus 8 reference strains (to help preserve gene nomenclature). In total, PIRATE identified 4,142 gene clusters, of which over half were characterized as core genes (present in 95% or more of the isolates; 2,150 genes; 52%). This was consistent with core genome estimates from other *S. aureus* collections (54–57). The accessory genome consisted of 1,992 genes (present in fewer than 95% of isolates), representing ∼48% of the pangenome. A large proportion of the accessory genome (78%) was present in fewer than 25% of isolates (1,57/1,992 accessory genes; **Supplementary table S3**). Core and accessory genome differences were visualized in phandango (58) (**Figure S1A**). Consistent with the clonal nature of *S. aureus*, isolates grouped by accessory genome content (using poppunk) clustered similarly to the clonal frame (**Figure S1BC**) (59).

#### Distribution of known virulence and AMR genes

All *S. aureus* isolates were sampled from invasive disease cases and contained many known virulence genes, with between 50 and 69 genes identified in each isolate from the VfDB database (average: 63; update March 2021). Nearly half of these putative virulence factors were present in all isolates (35 of 79; 44%), including the genes: *adsA* (phagocytosis escape), *aur* (metalloproteinase), *geh* (lipase), *hla (*α-hemolysin*), hlgAB (*γ*-*hemolysin), *hysA* (hyaluronate lysate), *icaABDR* (biofilm formation), *isdAB* (surface proteins), *lip* (lipase), *srtB* (surface protein anchor), *sspABC* (adhesion) and much of the *cap8* capsule operon (**Figure 2A**). Distribution of virulence genes between the two clinical outcome groups (“cured” and “not cured”) was also considered. The methicillin resistance gene, *mecA* was more prevalent in the “not cured” outcome group (n=2/21; 9.5%) compared to the “cured” group (n=3/65; 4.6%). Also, the presence of the *bbp* gene (bone sialprotein binding) (n=20/21; 95.2% versus n=58/65; 89.2%) and *ebpS* gene (elastin binding) (n=21/21; 100.0% versus n=60/65; 92.3%) were found to be more prevalent in the “not cured” outcome group than in the “cured” outcome group, but this was not statistically significant. (**Supplementary table S4**).

**Figure 2:**
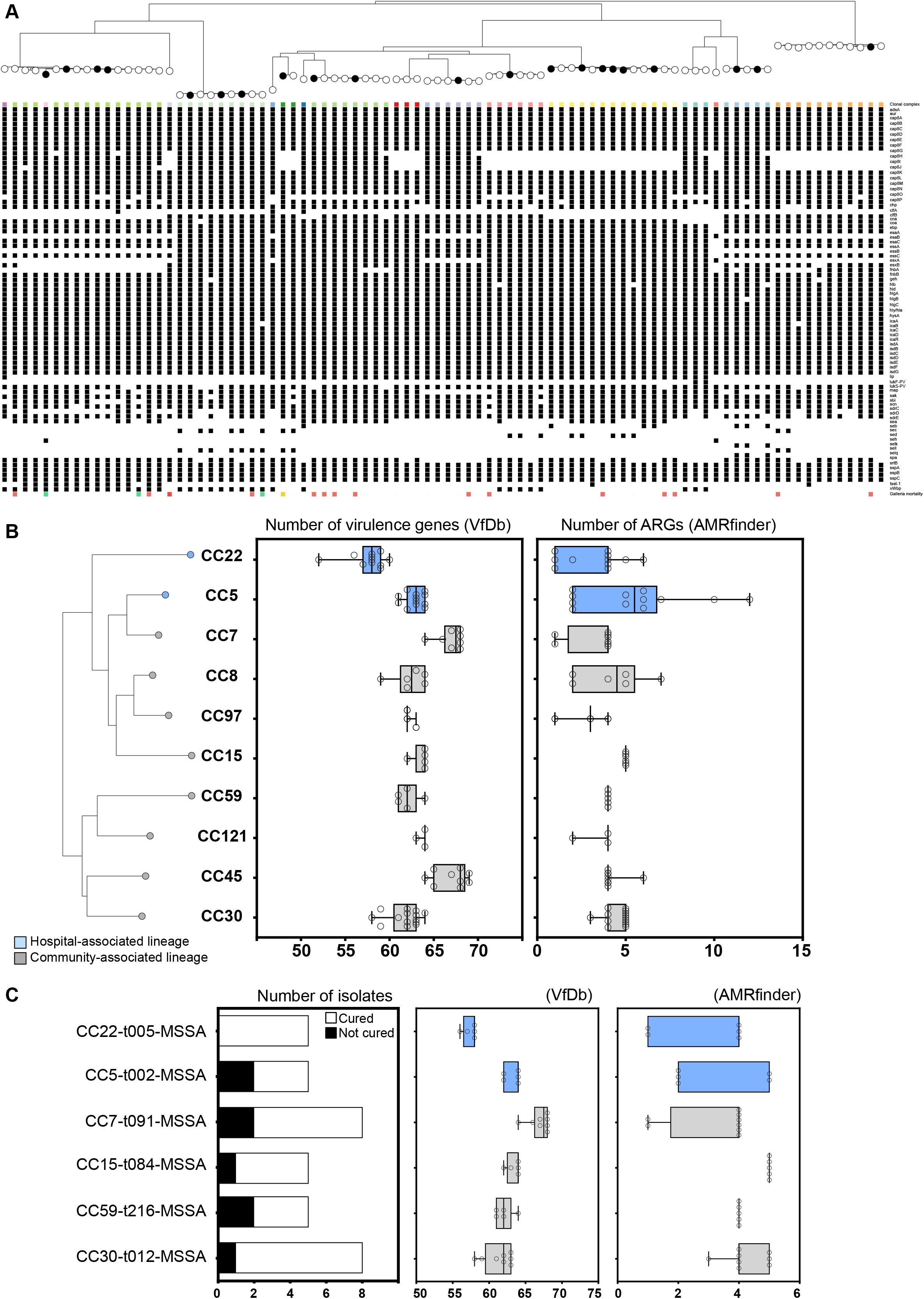
Hypervirulent lineages and clones. **A:** Maximum-likelihood phylogeny of our 86 *S. aureus* isolates above matrix of defined virulence factors identified using the VfDb (114). Summary boxplots of the number of virulence and antimicrobial resistance genes (ARGs) identified in each (**B)** clonal complex and (**C**) MSSA clone (represented by 3 or more isolates). All data points shown, bars show min and max. Hospital-associated lineages and clones highlighted in blue. The number of isolates from each MSSA clone is also shown, with the proportion of isolates from patients who did not achieve a” cured” status in black.

As most isolates were MSSA, there were very few antimicrobial resistance genes (ARGs) identified through nucleotide comparisons with the AMRfinder Plus database (**Supplementary table S5**). Differences were observed between clonal complexes for the numbers of virulence and AMR determinants (**Figure 2B**). In common with other studies, we identified slightly fewer virulence genes, but more ARGs in hospital-associated lineages. On average there were 60.3 virulence genes and 4.4 ARGs in HA lineages, compared with 63.8 virulence and 4.1 ARGs in community associated lineages. Six MSSA clones were represented by three or more isolates, of which five represented more than a third of our isolates (8 of 21; 38%) from patients who experienced a “not cured” outcome (**Figure 2C**). Two clones posed a particularly high risk to patients with 40% (2 of 5) of CC5-t002-MSSA and CC59-t216-MSSA isolates developing a “not cured” patient outcome. A quarter of patients (2 of 8) infected by CC7-t091-MSSA also experienced a “not cured” outcome, while those infected by the CC22-t005-MSSA clone all recovered.

#### Both “cured” and “not cured” isolates induced high mortality in a Galleria mellonella model

A selection of isolates (10 “not cured” and 10 “cured”) were used to challenge *G. mellonella* larvae. Bacterial suspensions were injected into larvae (average inoculum of 2.75×10^6^CFU/larvae; range: 1.87×10^6^ to 4.29× 10^6^) and incubated up to 120 hours. However, high mortality was observed in a proportion of isolates in both outcome groups (**Figure 3AB**). This may not be surprising as all isolates were from infection cases, and all were found to possess at least 12 toxin genes. Further analysis of the *Galleria* virulence results identified differences between the average mortality scores when specific putative virulence genes were present (**Figure 3C**). Differences in gene content between isolates were quantified and scored using SCOARY (60). We report minus log_10_ of the naïve p-value for the null hypothesis that the presence/absence of this gene is unrelated to the trait status (-log_10_>2 equivalent to naive p-value <0.01). We identified four gene clusters associated with increased killing during the *Galleria* infection model (**Figure 3D**). Three of which (g01257, g01221 and g01491) demonstrated more than 90% sequence similarity with a SOS response-activated pathogenicity island shared between *S. aureus* and *S. epidermidis* isolates (SACOL0900-0904) (54, 61). The fourth gene cluster (g03516) identified was identical (100% nucleotide similarity over 100% of the gene) to the *msaC* gene (SACOL1438/SAUSA300_1296). As an uncoded member of the *msaABCR* operon, this locus has an indirect role in virulence factors *aur, scp, ssp* and *spl* and contributes to increased virulence, biofilm formation and triggers the onset of bacterial persistence (62–64)(**Supplementary file S4**).

**Figure 3:**
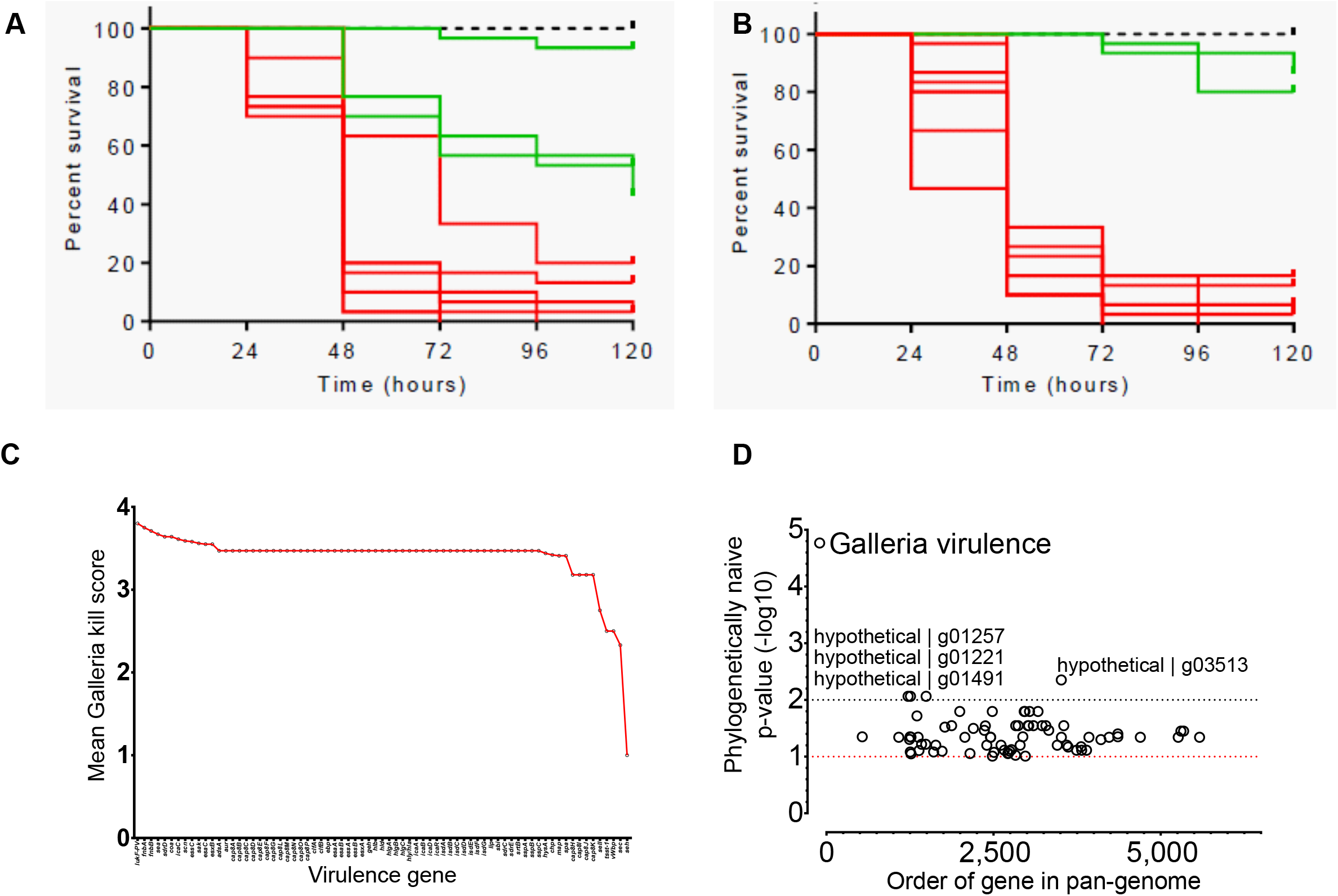
*Galleria mellonella* virulence model. Kaplan-Meier survival curve of larvae infected with *S. aureus* isolates from **A:** “not cured” outcome patients and from **B:** “cured” outcome patients. Each line represents a different *S. aureus* isolate injected into 10 larvae per isolate. Mean survival rate was calculated per isolate. Dotted line represents survival of larvae injected with PBS; green line represents high survival rate and red line indicates a high mortality rate. **C:** Mean kill curve scores from isolates that contained each virulence factor identified by VfDb. **D:** Pangenome-wide association study comparing isolates with high kill score (above 50% at 120 hrs) vs those with low kills scores using SCOARY (60). No lineage correction was used and the phylogenetically naïve minus LOG p-value reported. Three genes from previously identified pathogenicity island and *msaC* gene from the *msaABCR* operon were associated with increased killing.

#### Isolate phenotypic variation may contribute to the onset of infection, but does little to influence patient outcome

We tested laboratory phenotypes associated with increased virulence and compared these with patient outcome. Individually, none of the phenotypes contributed significantly to a “not cured” patient outcome (**Table 3**). Isolates were subject to antimicrobial susceptibility profiling (29 antibiotics), biofilm formation, hemolysis activity and staphyloxanthin production (**Supplementary table S6**). No isolate was considered extensively or pan drug resistant, although 4 were considered multi-drug resistant (resistant to three or more different classes of antibiotic). More than half (n=51/86; 59.3 %) of all collected isolates were unable to form a thick biofilm under laboratory conditions. Furthermore, 61.6 % (n=53/86) of all isolates produced staphyloxanthin and 39.5 % (n=34/86) showed hemolytic activity (**Table 3**). Isolates that demonstrated hemolytic activity contribute to a “cured” patient outcome (≥90% confidence; p=0.090, **Table 3**). Isolates that were able to produce a thick biofilm were often also able to produce staphyloxanthin (n=27/35; 77.1%; **Table 4**). However, poor biofilm forming isolates that produced staphyloxanthin (n=26/51; 51.0%) were detrimentally associated with a “not cured” patient outcome (n=15/26; 57.7%).

**Table 3:**
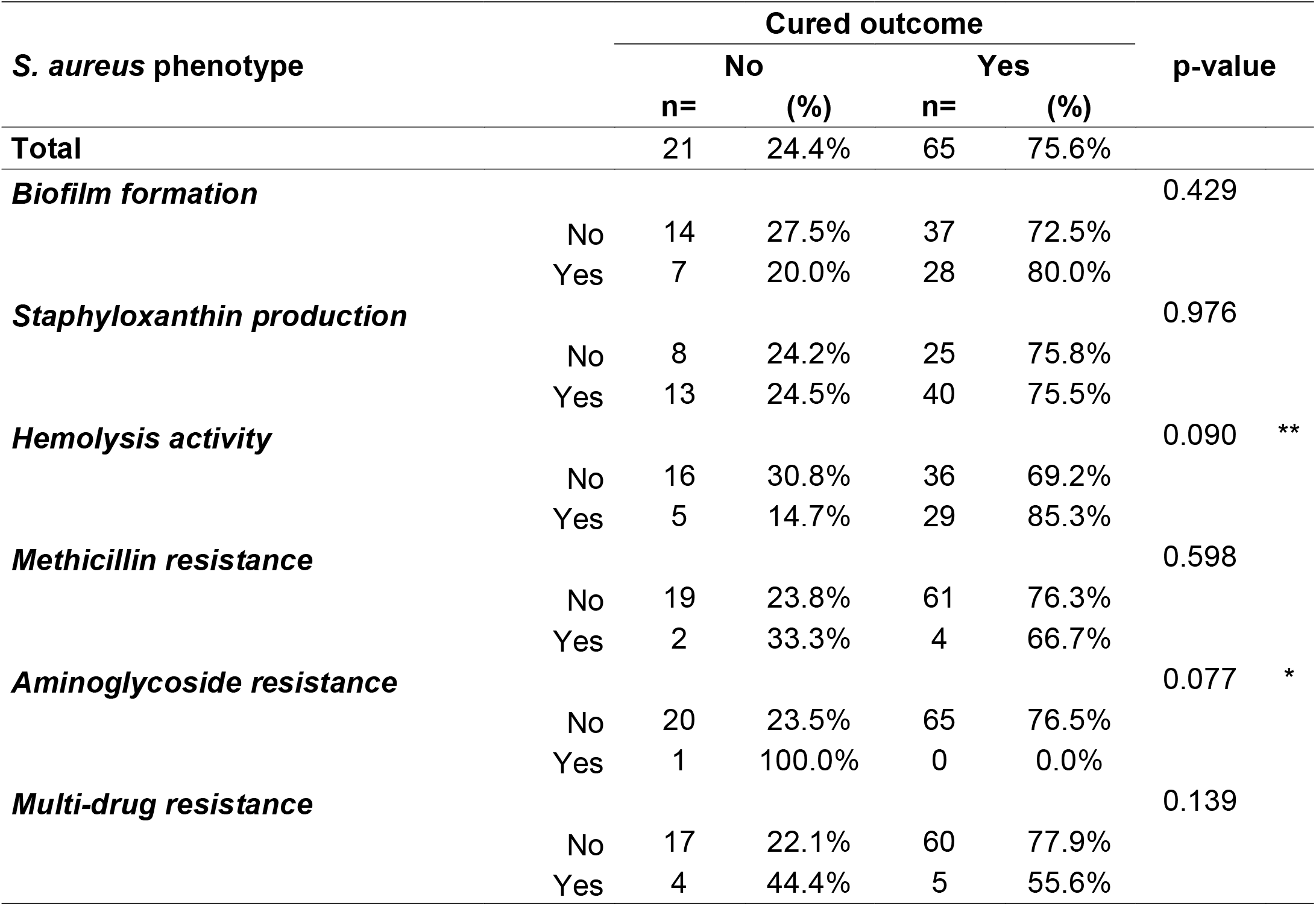
Phenotypic risk factors

**Table 4:**
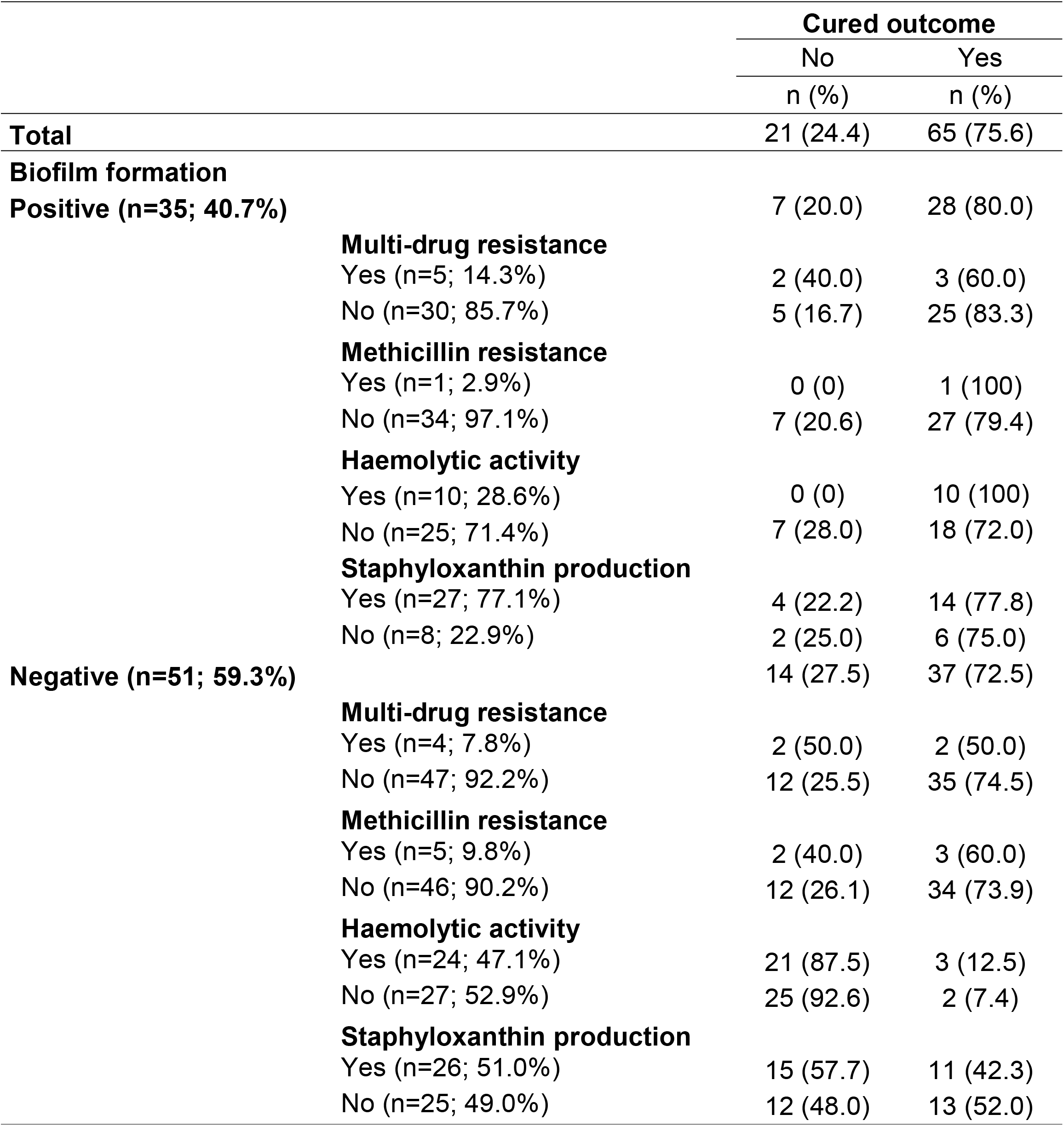
Phenotype associations in combination with enhanced biofilm formation.

#### Genes associated with different virulence phenotypes

Differences in gene content between phenotypes were quantified and scored using SCOARY, reporting the minus log_10_ of the naive p-value for the null hypothesis that the presence/absence of this gene is unrelated to the trait status (-log_10_>2 equivalent to naive p-value <0.01; **Supplementary table S7**). We identify genes associated with six different phenotypes, including patient outcome, biofilm formation, hemolysis, multi-drug resistance, methicillin resistance and staphyloxanthin production (**Figure 4)**. An uncharacterized membrane protein (g02811) and a gene from the SA97 virulent bacteriophage (g03902) were associated with a “not cured” patient outcome (-log_10_>2; naïve p-value <0.01; **Figure 4A**). Genes commonly found as part of the *SCCmec* cassette (*mecA, paaZ, upgQ* and *mecRI*) were associated with methicillin resistance, and were among the gene clusters that demonstrated the strongest association with any phenotype (-log_10_>6; naïve p-value <2*10^−7^; **Figure 4B**). Several hypothetical gene clusters were associated with biofilm formation (8 genes with naïve p-value <0.01; -log_10_>2; **Figure 4C**).

**Figure 4:**
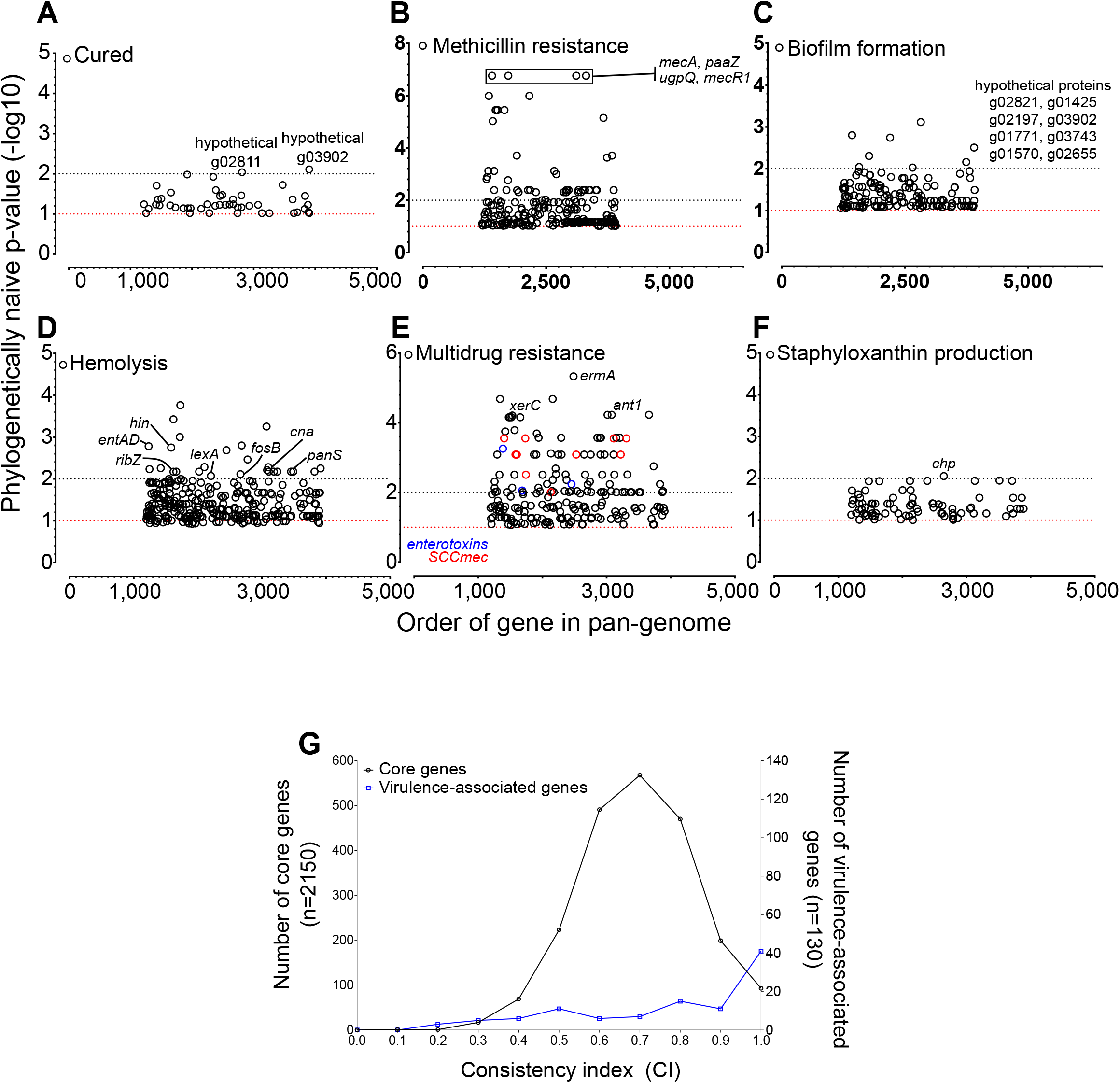
Pangenome-wide association studies. SCOARY was used to quantify gene presence between isolates tested for six targeted virulence phenotypes (60). No lineage correction was used, and the phylogenetically naïve minus LOG p-value reported. Isolates were scored for phenotypic differences in (**A**) patient outcome; (**B**) methicillin resistance; (**C**) biofilm formation; (**D**) hemolysis; (**E**) multidrug resistance; and (**F**) staphyloxanthin production. Genes that were highly associated with a given phenotype are labelled. Multiple toxin (blue circles) and *SCCmec* cassette (red circles) genes were associated with more than one phenotype. (**G**) Individual gene phylogenies were compared to a core phylogeny for each virulence phenotype-associated gene and consistency indices calculated. The two distributions were significantly different (Mann Whitney test; U = 78998, p < 0.0001) between virulence phenotype-associated genes (0.7766 ± 0.2418) compared with the average for all core genes (0.6943 ± 0.1408).

The presence of several genes (27 genes with -log_10_>2, lowest p-value: 0.00017), including enterotoxins A and D and the collagen adhesin *cna* were associated with hemolysis production (**Figure 4D**). Despite only four isolates that were classed as MDR, the presence of 93 genes (lowest p-value: 4.68×10^−6^) were associated with phenotypically measured MDR, which included several members of the *SCCmec* cassette, the erythromycin resistance-associated *ermA* and the aminoglycoside resistance determinant, *ant1* **Figure 4E**). Several enterotoxin genes were also associated with MDR, along with the biofilm-associated gene *xerC*. The chemotaxis inhibiting gene, *chp* was the only gene associated with staphyloxanthin production in our dataset (**Figure 4F**).

#### Genes linked to phenotypes that influence patient outcomes are more recombinogenic

Finally, we investigated the role of recombination in isolates that led to a poorer patient outcome. On average, isolates that led to a “not cured” patient outcome demonstrated the highest rates of recombination (**Figure S2**). However, genomes of isolates from patients who were successfully ‘cured’ were predicted to have included more recombination events. This is likely influenced by a small number of highly recombinogenic genomes that were predicted to have undergone extensive recombination **(Supplementary table S8**). Genes that were associated with any of our tested phenotypes demonstrated greater clonal inheritance than genes in the core genome. Meaning that phylogenies constructed from sequences of the individual genes were consistent with the phylogeny constructed from the core genome (**Figure 4G**). The mean consistency index (CI) was significantly higher (Mann Whitney test; U = 78998, p < 0.0001) among virulence phenotype-associated genes (0.7766 ± 0.2418) compared with the average for all core genes (0.6943 ± 0.1408). Taken together, the highly clonal population structure of *S. aureus* (long branches) – particularly those genes involved in virulence – and higher rates of recombination between lineages permits co-evolution of several successful hypervirulent lineages that can contribute to “not cured” patient outcomes in ODRI.

## Discussion

Despite advances in many aspects of emergency and orthopedic trauma care, ODRI persists as a challenge for the treating physicians and a significant burden for the patient. The interaction between the host immune system and invading isolates is complex and is not only affected by variation in host and bacterial genetics, but also by factors such as host wellbeing and general health (e.g. obesity). In a similar way to how the host deploys several different types of host defense mechanisms, bacteria are armed with a toolbox of different virulence factors. We used a combination of phenotype testing, an *in vivo* infection model and *in silico* genome characterization to identify lineages and virulence factors that may contribute to the risk of poor patient outcomes following ODRI.

Isolates from patients who experienced a “not cured” outcome did not cluster by core or accessory genomes (**Figure 1A**). The most common clonal complex associated with a poor patient outcome in our collection was CC5 (50%; **Figure 1B**). CC5 is highly recombinogenic and able to infect multiple host species (65, 66) and is often isolated from human infections, particularly skin and bone infections (55, 67). While infection is often caused by MRSA, there is growing concern for the spread of hypervirulent MSSA clones (67, 68). Nearly all the isolates we collected were sensitive to methicillin and lacked a functional *SCCmec* cassette (9 of 12 CC5 isolates). The gain and loss of *SCCmec* cassettes by multiple *S. aureus* clones continues to blur the differentiation of community-associated (CA) and hospital-associated (HA) *S. aureus* lineages (69, 70).

Since the emergence and epidemic spread of USA300 (CC8) clone in the early 2000s (30, 71, 72), CA-lineages have dominated infection surveillance studies (19, 71, 72). CA-lineages are typically thought to carry larger -more cumbersome, and less resistant -SCC*mec* types I, II and III, and often carry toxin genes such as the PVL genes. While traditional HA-*S. aureus* lineages carry smaller SCC*mec* cassettes that are more difficult to disrupt, which are maintained by successive generations. Several HA-lineages rose to prominence around the world, only to have been replaced as the most common lineages in human infections in specific regions; including the rise of CC22 in the UK and Europe (18, 73–75), and replacement of HA-MRSA-ST239 by CA-MRSA-ST59 in China (19). It is has been suggested that increased virulence, coupled with fewer putative AMR genes has contributed to the success of these CA-clones/lineages (31, 76). This was the case in our collection with more virulence genes and fewer ARGs found in CA-lineages compared to HA-lineages (**Figure 2B & Table S5**).

Isolates from the most common European lineage (CC22) were common among our collection of invasive isolates and were the most likely to lead to a “cured” patient outcome (**Figure 1B**). Further characterization of the specific clones in our collection identified 6 common clones, including a CC22-t005-MSSA clone that did not lead to a “not cured” patient outcome in any of the 5 patients it infected (**Figure 2C**). The five remaining clones that were identified in three of more patients were responsible for more than a third of the “not cured” patient outcomes. The CC59-t216-MSSA and CC5-t002-MSSA clones posed the highest risk, with 40% of patients infected by these clones developing a poor outcome. The CC5-t002-MRSA clone has been frequently observed, including studies from China and Iran where it exhibited extensive mupirocin resistance, MDR and contained several virulence toxins (PVL and tsst-1) (77, 78).

All isolates we collected were from invasive ODRI and contained at least 50 known virulence genes (**Figure 2AB**). Infection is complex and the function of any virulence gene is context dependent and a simple sum of the number of putative virulence elements is unlikely to be a good proxy for the infective potential of an isolate. All isolates in our collection were able to colonize and cause infection, but variation in additional virulence factors will influence disease progression and host evasion. Despite relatively small sample numbers, we were able to identify putative virulence-associated genes from an *in vivo* infection model and phenotypic assays, including biofilm production, antimicrobial susceptibility (and MDR), hemolysis and staphyloxanthin production.

Similar assays in *S. epidemidis* identified biofilm formation as key to establishing an ODRI through adhesion to implant surfaces (12, 43) and contributed towards a poorer clinical outcome, persistence and recurrence (8). However, this was not the case in the current study. Although 40% of the *S. aureus* isolates were able to form a strong biofilm, no clear influence on patient outcome was observed (**Table 3**). Armed with a suite of extracellular toxins, *S. aureus* can often cause more severe acute infections (79), while *S. epidermidis* is often associated with less severe, chronic infection – where strong biofilm formation on implants and dead tissue may be more relevant (80–82). *In vitro* Staphyloxanthin production leads to increased pathogenicity of *S. aureus* (83, 84), and 61% of our isolates were able to produce staphyloxanthin (**Table S6**). This level of prevalence is consistent with the observation by Post *et al*. whereby 56% of 305 isolates from implant and non-implant infections were staphyloxanthin producers (85). The chemotaxis inhibitor protein (CHIPS), encoded by *chp* was the only gene we found associated with staphyloxanthin production (**Figure 4G**). CHIPS ability to help evade host immune responses pose the potential for more severe and chronic infections (86, 87). Although it was not independently associated with “not cured” in our study population.

In hypervirulent MSSA lineages and clones, increased carriage of virulence genes is balanced against limited carriage of AMR genes. We observed this in our collection, with relatively few isolates resistant to multiple antibiotics (**Figure 2B & Table S5**), and lower rates of MRSA isolates compared to other studies (7% MRSA and 10% MDR). Rates of MRSA (27% & 24%) were much higher in other studies of ODRI (10, 11). Both these previous studies involved patients with a median age above 60, and patients infected with MRSA experienced lower cure rates (57%) than for MSSA-infected patients (72%) or CoNS (82%) (10). Very high rates of methicillin resistance have been observed in *S. epidermidis* ODRI isolates, which is consistent with different modes of infection between the species (73% MRSE and 76% MDR) (8). Elderly patients are more likely to suffer from infection with resistant bacteria due to repeated hospital stays and multi-morbidity (88, 89), and may partially explain the reduced number of MRSA isolates in our collection. Although only present in a small number of our isolates (n=6; 7%), methicillin resistance was associated with a “not cured” outcome (2 of 6; 33.3%; **Table 3**). This is consistent with many other studies that have highlighted the detrimental effect of methicillin resistance on treatment success (43, 55, 90).

Deep branching clades and highly structured clustering of isolates is evidence of the strong selection pressures that have driven the evolution of *S. aureus*. Distinct *S. aureus* lineages are found at markedly different prevalence globally, including hypervirulent sub-lineages (74, 91). Waves of pandemic sub-lineages have been described within CC30 - initially MSSA, before being replaced by MRSA variants (18, 19, 74). Independent acquisition of virulence determinants in these lineages was associated with the rise and fall of these lineages and horizontal gene transfer (HGT) likely has an important role in the accumulation of virulence factors. This lineage specific accumulation of virulence genes makes traditional identification of virulence factors difficult. Genome-wide association studies that can account for these lineage effects (43, 44, 90, 92), or can differentiate between the types of association, are potentially useful (93). Furthermore, the inclusion of the pangenome analyses (including mobile elements and intergenic regions) and covarying SNPs and genes (epistasis) (94, 95), combined with appropriate phenotypic validation and/or transcriptome profiling will greatly enhance understanding of the contribution of pathogen genetic variation on disease progression (49).

Infection is clearly a complex process influenced by the host immune system and genotypic variation within the invading pathogen. Here, we only tested a single *S. aureus* colony from each patient. However, there can be considerable diversity among the commensal and infective isolates within a single person (96–99). Small colony variants (SCV) (100, 101), persistence (102, 103) and dormancy (104, 105) also help protect invading cells from the host immune system and evade antimicrobial treatment. While the isolates collected here have already colonized host subcutaneous tissue and established an infection, our aim was to delineate factors that contribute to poorer patient outcomes. Our results are consistent with a complex balance between virulence and colonization (32), involving core and accessory genome elements, and provide some evidence that characterization of common emerging clones and lineages may help disease outcome prediction.

## Materials and methods

### Clinical data and Staphylococcus aureus collection

The clinical data and *S. aureus* collection was part of a prospective study performed between November 2011 and September 2013 at BGU Murnau in Germany (12) which was approved by the local ethical committee *Ethik-Kommission der Bayerischen Landesärztekammer* under approval number 12063 and registered with https://clinicaltrials.gov with identifier NCT02971657. All patients enrolled in the study were aged 18 or older and provided informed written consent. Patient inclusion criteria were culture-positive *S. aureus* infection involving fracture related infection (FRI) or periprosthetic joint infections (PJI). All patient details were anonymized, and *S. aureus* isolates given genome database identifiers (BIGSids), study identifiers (ARI-number) and sample laboratory identifiers (Lab-ID). Associated clinical data are summarized in **Table S1**. After an average of 23 months follow-up (FUP) patients were assessed for treatment outcome. When patients did not complete a follow-up appointment, the clinical outcome was assessed at the time of hospital discharge. “Cured” patients were free of infection, surgical and systemic antibiotic therapy had ceased with function of the affected joint or limb restored. If one or more of these parameters were negative, patients were considered to have had a “not cured” outcome (8, 12).

### Genome sequencing and assembly

*S. aureus* colonies were cultured in 5 ml Tryptone Soy Broth at 37 °C with overnight shaking. DNA was extracted using the QIAamp DNA Mini Kit (Qiagen, Germany) according to manufacturer’s instructions with the addition of 1.5 μg/μL lysostaphin (Sigma-Aldrich, Buchs, Switzerland) and 2 µg/ml lysozyme (Sigma-Aldrich, Buchs, Switzerland) to facilitate cell lysis. DNA was quantified using a spectrophotometer prior to sending for sequencing by Microsynth AG (Switzerland) using an Illumina MiSeq benchtop sequencer. Sequencing libraries were prepared using Nextera XT library preparation kits (v2) and paired end 250 bp reads generated with the MiSeq run kit (v2). Short read paired-end data was assembled *de novo* with SPAdes (version 3.3.0; using the *–careful* command)(106) and assessed for quality (n=86 genomes contained < 500 contigs). Average assembled genomes were 2,771,938 bp in length (range: 2,638,312 - 2,962,075 bp) consisting of 113 contigs (range: 29 – 358 contigs) with an N50 of 43,492 bp (range: 5,729 – 109,980) (**Table S2**).

### Genome archiving, multiple genome alignments and construction of isolate genealogies

An alignment of all 86 *S. aureus* isolates was constructed from concatenated gene sequences of all core genes (found in ≥95% isolates) using MAFFT (version 7)(107) on a gene-by-gene basis (size: 2,138,455 bp; **Supplementary file 1**). A maximum-likelihood phylogenies were constructed using a GTR + I + G substitution model and ultra-fast bootstrapping (1,000 bootstraps) implemented in IQ-TREE (version 2.0.3)(108, 109) and visualized on Microreact: https://microreact.org/project/Post-Pascoe-Saureus/0f0c26fb (**Supplementary file S3**) (110). Collection also shared on the pathogen watch website (https://pathogen.watch/collection/7rgjyrzz3xoc-post-and-pascoe-et-al-2022-hypervirulent-mssa-from-odri).

### Molecular typing and genome characterization

Our publicly available BIGSdb database (https://sheppardlab.com/resources/) includes functionality to determine multi-locus sequence type (MLST) profiles defined by the Staphylococcal pubMLST database (https://pubmlst.org/saureus; accessed March 2019) (111). Staphopia (version 1.0.0) was used to define *SCCmec* types (112) and *spa* types were typed *in silico* using spaTyper v1.0 (**Supplementary table S2**) (113).

### Care and accessory genome characterization

All unique genes present in at least one of our isolates (or eight reference strains) were identified by automated annotation using PROKKA (version 1.13) followed by PIRATE, a pangenomics tool that allows for orthologue gene clustering in bacteria (**Supplementary table S3; Supplementary file S3**). Genes families in PIRATE were defined using a wide range of amino acid percentage sequence identity thresholds for Markov Cluster algorithm (MCL) clustering (45, 50, 60, 70, 80, 90, 95, 98). Core genes were defined as present in 95% of the genomes and accessory genes as present in at least one isolate (**Supplementary figure S1A**). The pangenome was visualized using phandango, as a matrix of gene presence alongside a core genome phylogeny (58). Pairwise core and accessory genome distances were compared using PopPunk (version 1.1.4), which uses pairwise nucleotide k-mer comparisons to distinguish shared sequence and gene content to identify divergence of the accessory genome in relation to the core genome. A two-component Gaussian mixture model was used to construct a network to define clusters and visualized with microreact (**Supplementary figure S1BC**) (59, 110).

### Identification of known virulence traits and antimicrobial resistance genes

The presence of putative virulence factors and antimicrobial resistance genes were identified from assembled genomes using ABRICATE (v0.3) (59). Putative virulence genes were detected through comparison with reference nucleotide sequences in the VfDb (default settings: ≥70% identity over ≥50% of the gene; **Supplementary table S4**) (114). Antimicrobial resistance genes were identified using the NCBI AMRfinder Plus (115) and CARD (116) databases (19^th^ April, 2020 update). Results for both AMR databases were similar, with the CARD database identifying additional AMR associated genes. We report the results from the curated AMRfinder Plus database (**Supplementary table S5**).

### Phenotype testing

***Antibiotic susceptibility testing:*** Minimum inhibitory concentrations (MICs) for 29 antibiotics were determined using a Vitek2 machine (bioMérieux Vitek Inc., USA) as described previously (8, 12). Susceptibility breakpoints were defined according to the definitions of the European Committee of Antimicrobial Susceptibility Testing (EUCAST) and isolates resistant to 3 or more antimicrobial classes were defined as multidrug resistant (MDR)(118). The Vitek2 results for oxacillin and cefoxitin were used as a proxy for methicillin-resistance. ***Staphyloxanthin production:*** Staphyloxanthin production indicated by a yellow-orange pigmentation of *S. aureus* colonies as well as ***Haemolysin activity:*** Haemolysis activity of *S. aureus* strains was evaluated as previously described (119). ***Biofilm formation***: was also assayed as described previously (8, 12, 85, 120). Association amongst and between the clinical parameters, bacterial phenotypes, clades and presence/absence of genes were analyzed statistically using Chi-square test. Statistical analyses were performed using SPSS (Version 23, IBM, USA) or GraphPad prism 6 (GraphPad Software, Inc.; **Supplementary table S6**).

***In vivo virulence and survival assay in Galleria mellonella larvae:*** The invertebrate *G. mellonella* infection model was used to study the virulence of the *S. aureus* strains as previously described (121, 122). *G. mellonella* were obtained at pre-larval stage (Entomos AG, Zurich Switzerland). Larvae were grown at 30 °C in the dark and groups of ten larvae in the final instar larval stage weighing 200– 400 mg were used in all assays. A bacterial suspension of 10^6^ CFU/ml was prepared. Quantitative culture of a sample from each bacterial suspension was performed immediately after preparation by ten-fold serial dilution and plating on Tryptone Soy Agar plates to check the actual total viable count of the prepared suspension. Bacterial inoculates (10 µl) were injected into the last left pro-leg into the hemocoel of the last-instar larvae (200-400 mg). After injection, larvae were incubated at 37 °C in the dark. Larvae were assessed daily for survival up to 5 days post-injection and were evaluated according to survival, being scored as dead when they displayed no movement in response to touch. Controls included a group of larvae that did not receive any injection and a group of larvae inoculated with sterile phosphate-buffered saline (PBS). Experiments consisted of 10 larvae per bacterial strain, which was repeated in three separate experiments. For *G. mellonella* survival analysis, larvae mean survival curves were plotted using the Kaplan-Meier method (GraphPad Prism 6, USA; **Supplementary table S6**).

### Pan-genome-wide association studies of phenotype variation

Pangenome-wide differences in gene presence were quantified using the genome-wide association software, SCOARY (version 1.6.14)(60). With only limited numbers of samples in our collection, we report phylogenetically naïve differences in gene presence between clinical and laboratory symptoms and phenotypes (**Supplementary table S7**).

### Recombination and allelic consistency

The number of polymorphisms introduced by mutation and recombination in the core genome were inferred using Gubbins (version 2.4.1)(123) for each isolate (per branch; **Supplementary table S8**).The consistency of the phylogenetic tree to patterns of variation in sequence alignments for each gene of interest was calculated as before (124, 125). Consistency indices for each single-gene alignment of 130 virulence-associated genes to a phylogeny constructed using an alignment of 2,150 core genes shared by 94 isolates, were calculated using the CI function of the R Phangorn package (126).

## Data Availability

Illumina short read sequence data are archived on the Sequence Read Archive. All assembled genomes are shared on figshare (doi.org: 10.6084/m9.figshare.7926866) and are available on our public Staphylococcal Bacterial Isolate Genome Sequence Database (BIGSdb): https://sheppardlab.com/resources/. Collection also shared on the pathogen watch website (https://pathogen.watch/collection/7rgjyrzz3xoc-post-and-pascoe-et-al-2022-hypervirulent-mssa-from-odri). Isolate genealogy with associated meta-data is visualized on microreact [110]: https://microreact.org/project/Post-Pascoe-Saureus/0f0c26fb.

## Data availability

Illumina short read sequence data are archived on the Sequence Read Archive associated with BioProject accession PRJNA529795. All assembled genomes are shared on figshare (doi: 10.6084/m9.figshare.7926866) and are available on our public Staphylococcal Bacterial Isolate Genome Sequence Database (BIGSdb):
https://sheppardlab.com/resources/. Isolate genealogy with associated meta-data is visualized on microreact (110): https://microreact.org/project/Post-Pascoe-Saureus/0f0c26fb. All high-performance computing was performed on MRC CLIMB (127) in a bioconda environment (128).

## Acknowledgements

This work was funded by AO Trauma as part of the Clinical Priority Program, Bone Infection. All high-performance computing was performed on MRC CLIMB, funded by the Medical Research Council (MR/L015080/1 & MR/T030062/1). This publication made use of the PubMLST website (http://pubmlst.org/) developed by Keith Jolley and Martin Maiden (129) and sited at the University of Oxford. The development of that website was funded by the Wellcome Trust. The funders had no role in study design, data collection and interpretation, or the decision to submit the work for publication.

All authors report no conflicts of interest relevant to this article.

## Contributions

Conceptualization: VP, BP, SKS, and TFM; Isolate collection: VP, CE, JF, MM, and TFM; Data collection and curation: VP, BP, MDH and SKS; Formal Analysis: VP, BP, EM, JKC; Visualization: VP, BP, EM, JKC; Writing – original draft: VP, BP, SKS and TFM; Writing – Review & Editing: VP, BP, SKS and TFM; Funding Acquisition: VP, RGR and TFM.

## Figures and Table legends

**Table 1:** Collection overview.

**Table 2:** Patient risk factors.

**Table 3:** Infection isolate prognostic phenotypes.

**Table 4:** Phenotype associations in combination with enhanced biofilm formation.

**Supplementary material (**figshare: https://doi.org/10.6084/m9.figshare.7926866)

**Table S1:** Demographic data collected for each isolate.

**Table S2:** Genome quality control and extended typing results.

**Table S3:** Summary of core and accessory genome characterization with PIRATE.

**Table S4:** Summary of genes identified by screening known virulence genes with the VfDB.

**Table S5:** Summary of genes identified by screening antibiotic resistance genes with the AMRfinder (NCBI) and CARD databases.

**Table S6:** Summary of virulence phenotype data.

**Table S7:** Summary of genes associated with virulence phenotypes (SCOARY).

**Table S8:** Per isolate recombination statistics using Gubbins.

**Figure S1:** Accessory genome characterization.

**Figure S2**: Recombination analysis.

**Supplementary file 1:** Contigs

**Supplementary file 2:** Alignment.

**Supplementary file 3:** Phylogeny (.nwk) of 86 collected isolates.

**Supplementary file 4**: PIRATE pangenome gene families.

