## Supplementary figures and images for "Multiple hypervirulent methicillin-sensitive *Staphylococcus aureus* lineages contribute towards poor patient outcomes in orthopedic device-related infections"

### Figure S1

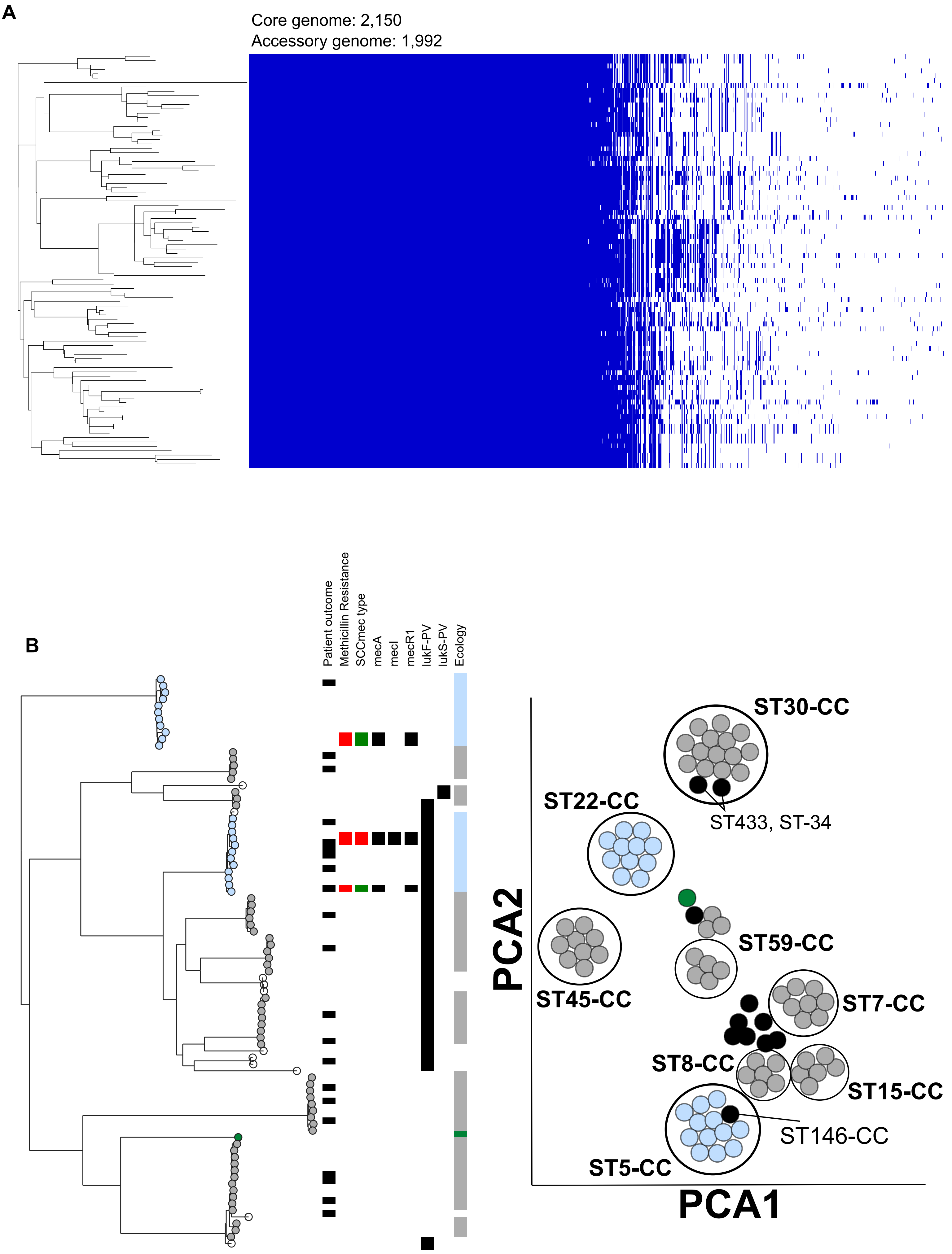

### Figure S2

A

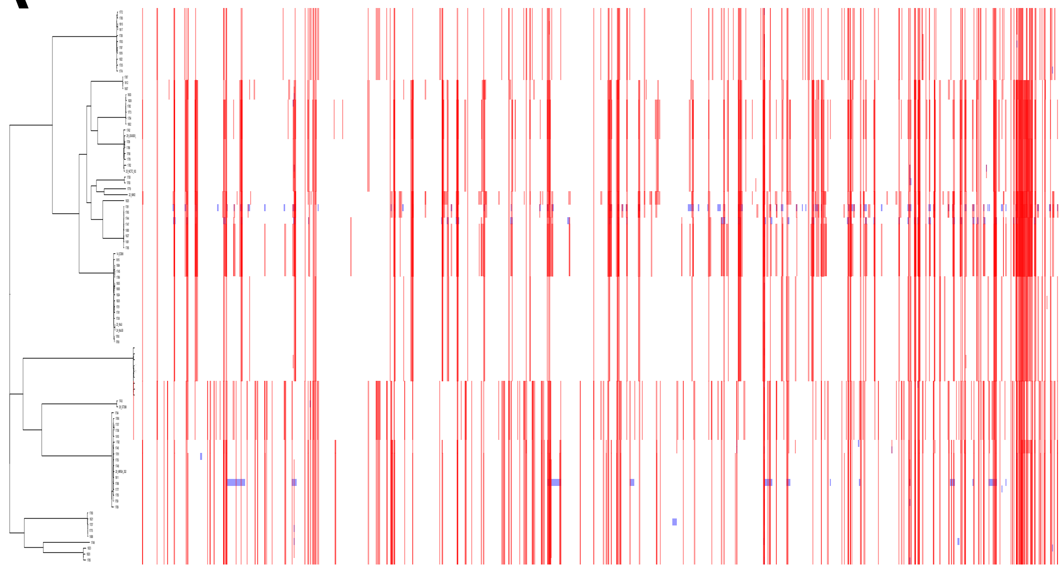

B

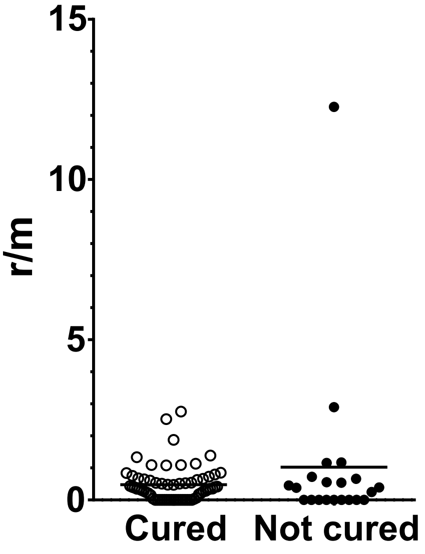
